# Active-day Exercise Duration and Cardiovascular Health Metrics: Novel Insights from NHANES 2007 – 2020

**DOI:** 10.1101/2025.09.29.25336932

**Authors:** Xiaodong Shang, Hengwei Zhang, Jinxiao Hou, John Oruongo, Cheng Cheng, Gongping Sun, Jin Liang, Qian Jun

## Abstract

**Background:** Current physical activity (PA) guidelines emphasize weekly volume, but how activity is distributed across active days may also influence cardiovascular health (CVH). We examined whether these weekly accumulation patterns are associated with CVH.

**Methods:** We conducted a cross-sectional analysis of adults aged 20–64 years without established cardiovascular disease (CVD) in the National Health and Nutrition Examination Survey (2007- March 2020). PA was self-reported and categorized as inactive (< 30 min/week), low-volume (30–89 min/week), intermediate-volume (90–149 min/week), or guideline-adherent (≥150 min/week). Within low- and intermediate-volume groups, participants were further classified as distributed (<30 or < 45 minutes on active days) or concentrated (≥30 or ≥ 45 minutes on active days). CVH was assessed using the American Heart Association’s Life’s Essential 8 (LE8). Associations were examined with LOESS smoothing and propensity score matching.

**Results:** Among 9,851 participants, 48.7% were inactive and only 33.2% met guideline PA levels. In the low-volume group, individuals with concentrated patterns (≥30 minutes on active days) had significantly better blood pressure scores compared with those with distributed patterns (<30 minutes; p=0.045). In the intermediate-volume group, the concentrated patterns (≥45 minutes on active days) were associated with more favorable glycemic (p<0.001) and lipid profiles (p=0.013), independent of total weekly volume. Dose-response analyses identified 30–45 minutes on active days as the optimal range for cardiometabolic benefits.

**Conclusion:** Weekly accumulation patterns independently influenced CVH among adults below guideline PA levels. Concentrating ≥ 30-45 minutes on active days was associated with better blood pressure, glucose, and lipid profiles without increasing weekly volume. These findings highlight active-day duration as a novel, modifiable, and time-efficient behavioral target for cardiovascular prevention.

**CLINICAL PERSPECTIVE:** *What is New?:*

- This is the first study to show that the way activity is structured across active days— whether spread in shorter bouts or sustain longer sessions —independently affects cardiovascular health.
- Accumulating ≥30–45 minutes of activity on active days was linked to better blood pressure, glucose, and lipid profiles compared with shorter distributed durations, even without increasing total weekly minutes.

*What are the Clinical Implications?:*

- Clinicians can shift counseling from simply "do more" toward "do smarter": how patients distribute their activity matters for cardiovascular health.
- Recommending ≥30–45 minutes of sustained activity on active days offers an efficient, achievable strategy that benefits patients who cannot meet weekly guideline totals.
- Framing active-day duration as a time-efficient and equitable behavioral target provides a practical pathway for personalized cardiovascular prevention.

## INTRODUCTION

Cardiovascular disease (CVD) remains the leading cause of death in the United States and worldwide, accounting for nearly one-third of all global mortality^1,2^ and imposing an annual economic burden exceeding $400 billion in the US alone.^3^ Despite major advances in pharmacologic and interventional therapies, prevention through lifestyle modification remains the most cost-effective and sustainable strategy for reducing cardiovascular risk.^4–6^

To standardize the measurement of cardiovascular health (CVH), the American Heart Association (AHA) introduced Life’s Essential 8 (LE8) in 2022, building on the earlier Life’s Simple 7 framework.^7,8^ LE8 includes four health behaviors—diet, physical activity (PA), nicotine exposure, and sleep—and four measurable health factors—body mass index (BMI), blood lipids, blood glucose, and blood pressure. By explicitly recognizing that upstream behaviors shape downstream biological measures, LE8 highlights the strategic importance of optimizing lifestyle interventions at both the individual and population levels.^8^

Among these behaviors, PA is one of the most powerful determinants of CVH. Current guidelines recommend a minimum of 150 minutes of moderate-intensity or 75 minutes of vigorous-intensity activity per week.^9,10^ While the benefits of meeting these thresholds are well established,^11,12^ less is known about how activity is distributed across the week. Some individuals exercise briefly on many days, while others concentrate longer durations into fewer active days. These weekly accumulation patterns may be an overlooked dimension of CVH.

Experimental studies provide biological plausibility for why the duration of activity on active days may matter independently of total weekly minutes. Sustained activity of at least 20-30 minutes can lower blood pressure for up to 24 hours,^13,14^ enhance glucose uptake through GLUT4 translocation,^15^ and stimulate mitochondrial adaptations^16^—effects unlikely to be triggered by very short activity periods. Collectively, these findings suggest that concentrating activity into longer durations on active days could yield cardiovascular benefits, particularly among adults who fall short of guideline-recommended totals.

To address this question, we leveraged data from the National Health and Nutrition Examination Survey (NHANES) 2007 - March 2020. Using the self-reported PA questionnaire, which inquired about time spent in moderate-to-vigorous activity on a typical day, we classified adults below guideline levels into distributed or concentrated weekly accumulation patterns based on their reported active-day duration. We then evaluated associations with CVH using the AHA LE8 framework. This population-based analysis provides new insight into whether the organization of activity across the week represents a practical and time-efficient target for cardiovascular prevention.

## METHODS

### Study Design and Population

We conducted a cross-sectional analysis using data from the NHANES 2007- March 2020, which included five complete two-year cycles (2007-2016) and one pre-pandemic period (2017- March 2020). We excluded the 2021-2023 cycle to avoid potential bias from pandemic-related changes in PA, social behaviors, and healthcare access.^17^ NHANES employs a complex, multistage probability sampling design to represent the non-institutionalized US population.

Of 66,148 participants examined, we restricted the analytic cohort to adults aged 20-64 years without established cardiovascular disease (CVD), as primary prevention is most relevant in this group. We excluded participants who were pregnant (n=404), current breastfeeding (n=299), or with self-reported CVD (congestive heart failure [n=482], coronary heart disease [n=523], angina pectoris [n=413], myocardial infarction [n=637], or stroke [n=637]). We also excluded those with malignancy (n=1,489) due to potential cardiometabolic effects of cancer therapies. Participants with incomplete LE8 data (n=15,476) were also removed, yielding a final analytic cohort of 9,851 individuals (*Supplementary Figure s1*).

Demographic variables (age, sex, and race/ethnicity) were self-reported. Race/ethnicity categories included non-Hispanic Asian, non-Hispanic Black, Mexican American, other Hispanic, non-Hispanic White, and multiracial/other. Estimates for non-Hispanic Asian participants became available beginning in the 2011-2012 cycle.

### PA Assessment

PA was assessed using the NHANES questionnaire, which asked participants to report frequency (days per week) and duration (minutes on a typical day) separately for moderate-intensity (small increases in breathing/heart rate) and vigorous-intensity (large increases) activities. Weekly PA volume was calculated by multiplying weekly frequency by reported active-day duration and summing across moderate and vigorous components.

We stratified participants by weekly PA volume: inactive (< 30 minutes/week), low-volume (30-89 minutes/week), intermediate-volume (90-149 minutes/week), and guideline-adherent (≥ 150 minutes/week). Within the low- and intermediate-volume groups, we further classified participants by weekly accumulation pattern. The distributed group included those reporting <30 minutes on active days in the low-volume group or <45 minutes on active days in the intermediate-volume group. The concentrated group included those reporting ≥30 or ≥45 minutes on active days, respectively. These thresholds approximated within-group medians and corresponded to physiological transitions in exercise response.

### CVH Assessment

CVH was operationalized using the AHA’s LE8 framework. Each of the eight components was scored on a 0-100 scale, with higher scores indicating more favorable health.^18^ Behaviors included diet (DASH-style score from two 24-hour dietary recalls^19^), PA (active-day duration and frequency as above), nicotine exposure (cigarette use, electronic nicotine delivery systems, secondhand smoke), and sleep (self-reported hours per night). Health factors included BMI (measured), blood lipids (laboratory total and HDL cholesterol, with non-HDL cholesterol calculated), glycemic control (fasting glucose or HbA1c), and blood pressure (average of three standardized seated measurements).

### Statistical Analysis

All primary analyses were conducted within the analytic cohort of 9,851 adults. Continuous variables were summarized as medians with interquartile ranges (IQRs), and categorical variables as proportions. Between-group differences were assessed using Mann-Whitney U tests for continuous outcomes and chi-square (χ^2^) tests for categorical outcomes, a standard approach for non-normally distributed data.^20^

Dose-response relationships between active-day duration (10-60 minutes) and LE8 health factors were explored using locally weighted scatterplot smoothing (LOESS, span=0.75) with 95% confidence intervals.^21^ Subgroups with <10 participants were excluded, and outliers were removed using a robust rule (±1.5 IQR from the median).

To minimize confounding, we performed 1:1 propensity score matching (PSM) within low- and intermediate-volume groups to compare distributed versus concentrated patterns. Propensity scores were estimated using logistic regression, including age, sex, race/ethnicity, education, income-to-poverty ratio, and non-PA health behaviors (diet, smoking, sleep). ^22,23^ Nearest neighbor matching with a caliper width of 0.05 standard deviations of the logit of the propensity score was applied.^24^ Covariate balance was evaluated using standardized mean differences (SMDs), with |SMD| < 0.10 considered acceptable.^25^ Post-matching comparisons used Wilcoxon signed-rank tests for continuous outcomes and Stuart-Maxwell test^26,27^ for categorical variables with ≥ 3 levels, preserving matched-pair structure.

For sensitivity analyses, we repeated survey-weighted descriptive statistics and regression models incorporating NHANES sampling weights, strata, and primary sampling units to generate nationally representative estimates.^28–30^ These results were directionally consistent with the analytic cohort and are presented in the Appendix.

All analyses were conducted in Python 3.11.11, using pandas (v2.2.2), statsmodels (v0.14.4), and scikit-learn (v1.6.1). Two-sided P < 0.05 is defined as statistically significant. No additional adjustments were made for multiple comparisons in the primary analysis.

## RESULTS

The final analytical cohort comprised 9,851 US adults aged 20-64 years without established CVD (*Table 1*). The sample was evenly distributed by sex (50.4% male, 49.6% female), with similar median ages (41 years for men, 42 years for women). More than half of the participants were aged 40-64 years. Non-Hispanic Whites comprised the largest racial group, followed by Hispanic and non-Hispanic Blacks. Educational attainment and income-to-poverty ratio differed by sex: men were more likely to report a high school education or less, whereas women were more likely to have completed college. Men reported higher household income relative to the poverty level.

**Table 1.**
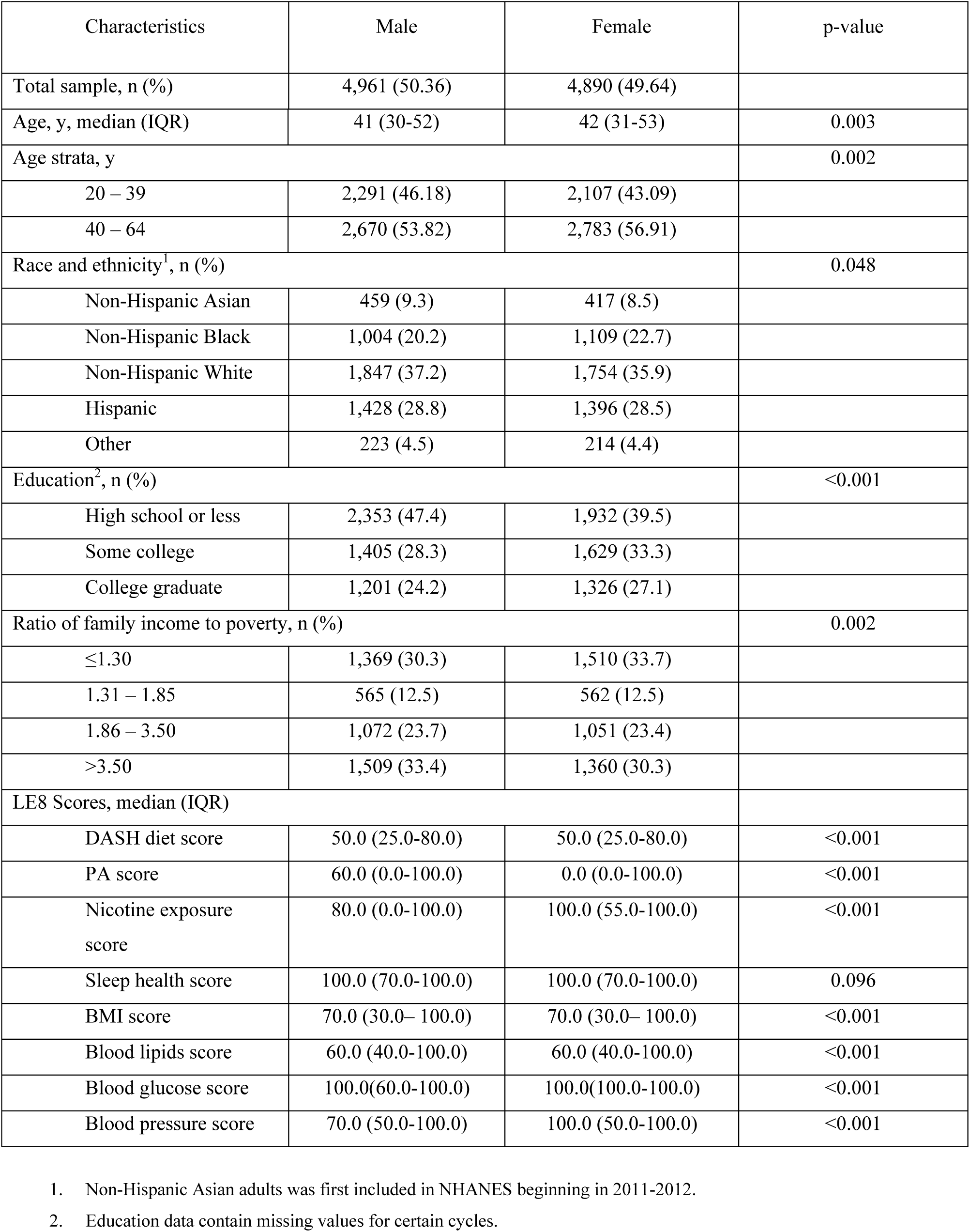
Baseline characteristics of the analytic cohort by sex (NHANES 2007 – March 2020).

| Characteristics | Male | Female | p-value |
| --- | --- | --- | --- |
| Total sample, n (%) | 4,961 (50.36) | 4,890 (49.64) |  |
| Age, y, median (IQR) | 41 (30-52) | 42 (31-53) | 0.003 |
| Age strata, y |  |  | 0.002 |
| 20 – 39 | 2,291 (46.18) | 2,107 (43.09) |  |
| 40 – 64 | 2,670 (53.82) | 2,783 (56.91) |  |
| Race and ethnicity<sup>1</sup>, n (%) |  |  | 0.048 |
| Non-Hispanic Asian | 459 (9.3) | 417 (8.5) |  |
| Non-Hispanic Black | 1,004 (20.2) | 1,109 (22.7) |  |
| Non-Hispanic White | 1,847 (37.2) | 1,754 (35.9) |  |
| Hispanic | 1,428 (28.8) | 1,396 (28.5) |  |
| Other | 223 (4.5) | 214 (4.4) |  |
| Education<sup>2</sup>, n (%) |  |  | <0.001 |
| High school or less | 2,353 (47.4) | 1,932 (39.5) |  |
| Some college | 1,405 (28.3) | 1,629 (33.3) |  |
| College graduate | 1,201 (24.2) | 1,326 (27.1) |  |
| Ratio of family income to poverty, n (%) |  |  | 0.002 |
| ≤1.30 | 1,369 (30.3) | 1,510 (33.7) |  |
| 1.31 – 1.85 | 565 (12.5) | 562 (12.5) |  |
| 1.86 – 3.50 | 1,072 (23.7) | 1,051 (23.4) |  |
| >3.50 | 1,509 (33.4) | 1,360 (30.3) |  |
| LE8 Scores, median (IQR) |  |  |  |
| DASH diet score | 50.0 (25.0-80.0) | 50.0 (25.0-80.0) | <0.001 |
| PA score | 60.0 (0.0-100.0) | 0.0 (0.0-100.0) | <0.001 |
| Nicotine exposure score | 80.0 (0.0-100.0) | 100.0 (55.0-100.0) | <0.001 |
| Sleep health score | 100.0 (70.0-100.0) | 100.0 (70.0-100.0) | 0.096 |
| BMI score | 70.0 (30.0– 100.0) | 70.0 (30.0– 100.0) | <0.001 |
| Blood lipids score | 60.0 (40.0-100.0) | 60.0 (40.0-100.0) | <0.001 |
| Blood glucose score | 100.0(60.0-100.0) | 100.0(100.0-100.0) | <0.001 |
| Blood pressure score | 70.0 (50.0-100.0) | 100.0 (50.0-100.0) | <0.001 |

1. Non-Hispanic Asian adults was first included in NHANES beginning in 2011-2012. 2. Education data contain missing values for certain cycles.

Baseline LE8 metrics also varied by sex. Women demonstrated more favorable scores for nicotine exposure, blood glucose, and blood pressure, while men had higher scores for PA and BMI (*Table 1*). These differences highlight sex-specific variation in CVH profiles.

### Prevalence and Distribution of PA Patterns

Nearly half of participants (48.7%) were classified as inactive (<30 min/week), while only one-third (33.2%) met current PA guidelines (≥150 min/week) (*Table 2*). Among those not meeting guidelines but reporting structured activity, 8.0% were classified as low-volume PA (30-89 min/week) and 10.1% as intermediate-volume (90-149 min/week). Women had higher rates of inactivity and lower rates of guideline adherence compared with men (*Table 2*).

**Table 2.**
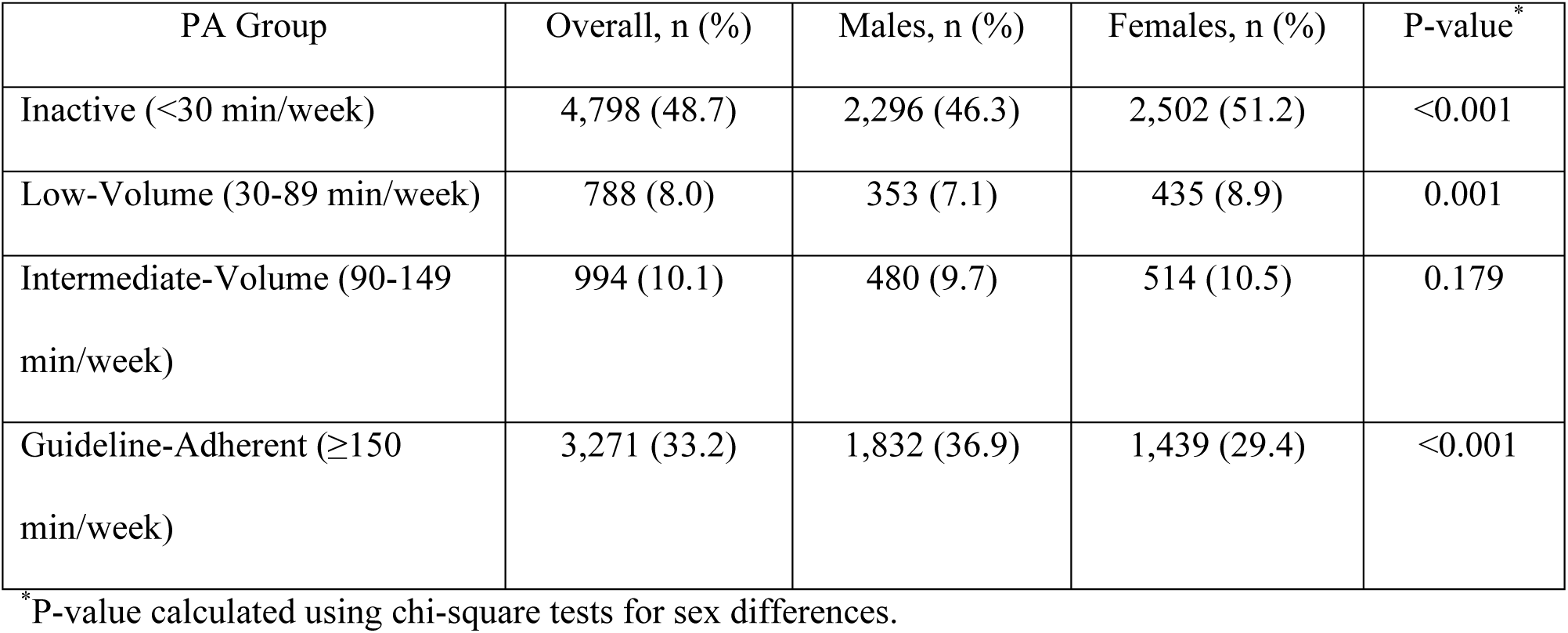
Distribution of PA categories in the analytic cohort, overall and by sex.

### Association Between PA Categories and LE8 Metrics

LE8 component scores improved stepwise with higher weekly PA volume (*Figure 1*). Inactive participants had the lowest scores across nearly all domains, particularly nicotine exposure and blood glucose. Among inactive adults, the 25^th^ percentile (Q1) nicotine score was 0, and the median was 80. In contrast, in the low-, intermediate-, and guideline-adherent groups, the 25^th^ percentile was ≥50, and the median was 100. Low-volume activity was associated with modest improvements, intermediate-volume participants showed further gains in blood pressure and lipid profiles, and guideline-adherent individuals had the most favorable scores across all domains. Diet scores remained consistently low, and sleep health scores consistently high, regardless of PA category.

**Figure 1.**
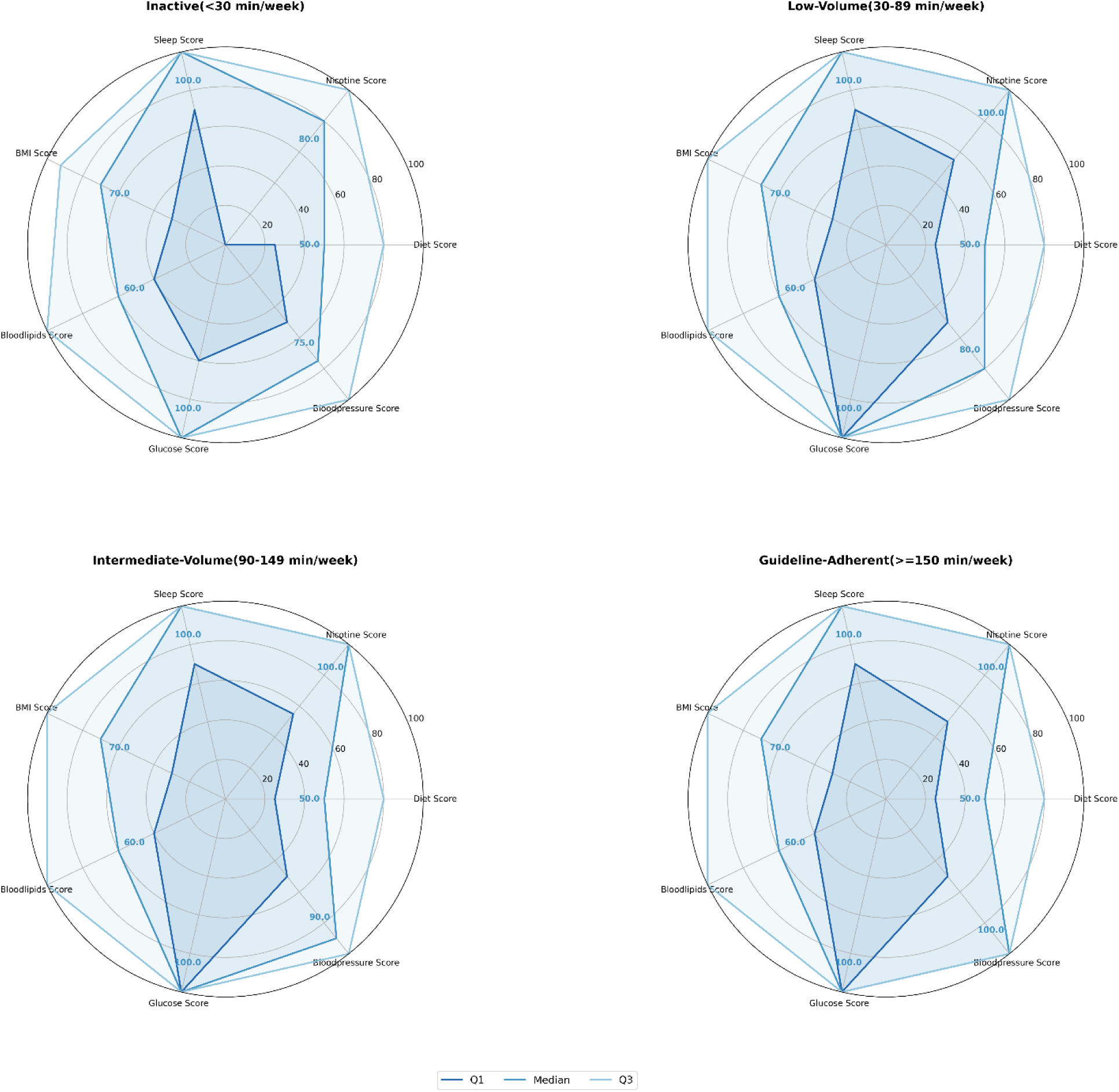
LE8 component scores by PA category. Radar plots display median values (solid lines) and interquartile range (shaded area) of LE8 scores (0–100 scale) for inactive, low-volume, intermediate-volume, and guideline-adherent groups.

### Dose-response of Active-Day Duration

Active-day duration was associated with cardiometabolic health among adults performing <150 min/week of PA (*Figure 2*). In the low-volume group, accumulating 30–45 minutes on active days was associated with higher BMI, blood pressure, and glucose scores. For example, blood pressure scores improved from 70 at shorter durations to near 95 at 45 minutes. In the intermediate-volume group, optimal scores for BMI, glucose, and lipids generally occurred at 45–50 minutes, although estimates were less stable at the extremes.

**Figure 2.**
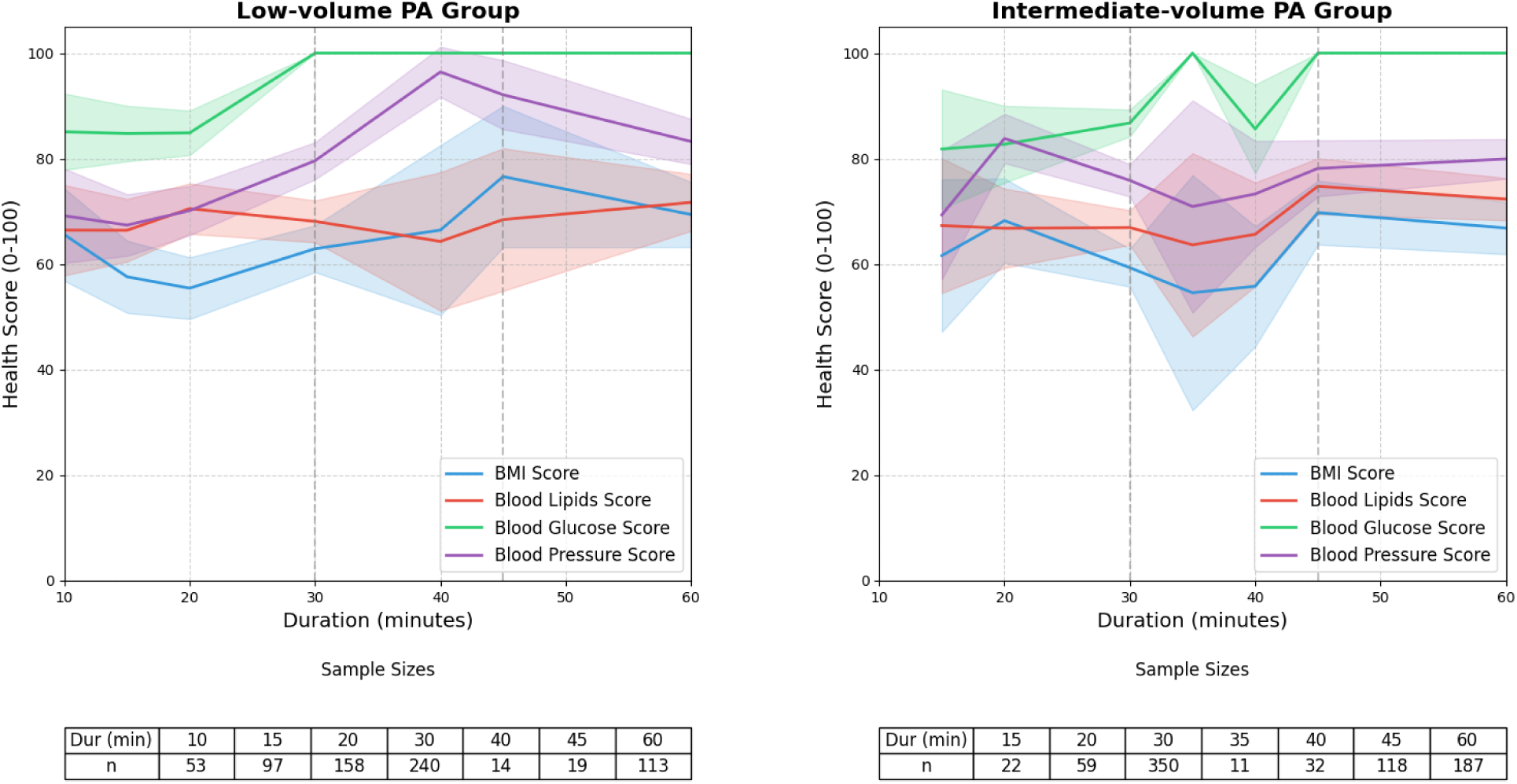
Active-day duration and LE8 health factors. LOESS curves with 95% confidence bands for BMI, blood lipids, glucose, and blood pressure by active-day duration. Panel A: low-volume (30–89 min/week). Panel B: intermediate-volume (90–149 min/week).

### Matched Comparisons by Weekly Pattern

#### Low-Volume Stratum

Among 788 low-volume participants, 40.2% reported distributed (<30 minutes on active days) and 59.8% concentrated (≥30 minutes). Before matching, demographic imbalances were observed, with concentrated exercisers more likely to be younger and female. After PSM, 249 matched pairs achieved excellent covariate balance (all |SMD|s <0.05; *Table 3A*). Concentrated patterns were associated with significantly higher blood pressure scores compared with distributed patterns (median 100 vs. 75; p=0.045; *Table 4A, Figure 3A*), while BMI, glucose, and lipid scores were not significantly different.

**Figure 3A.**
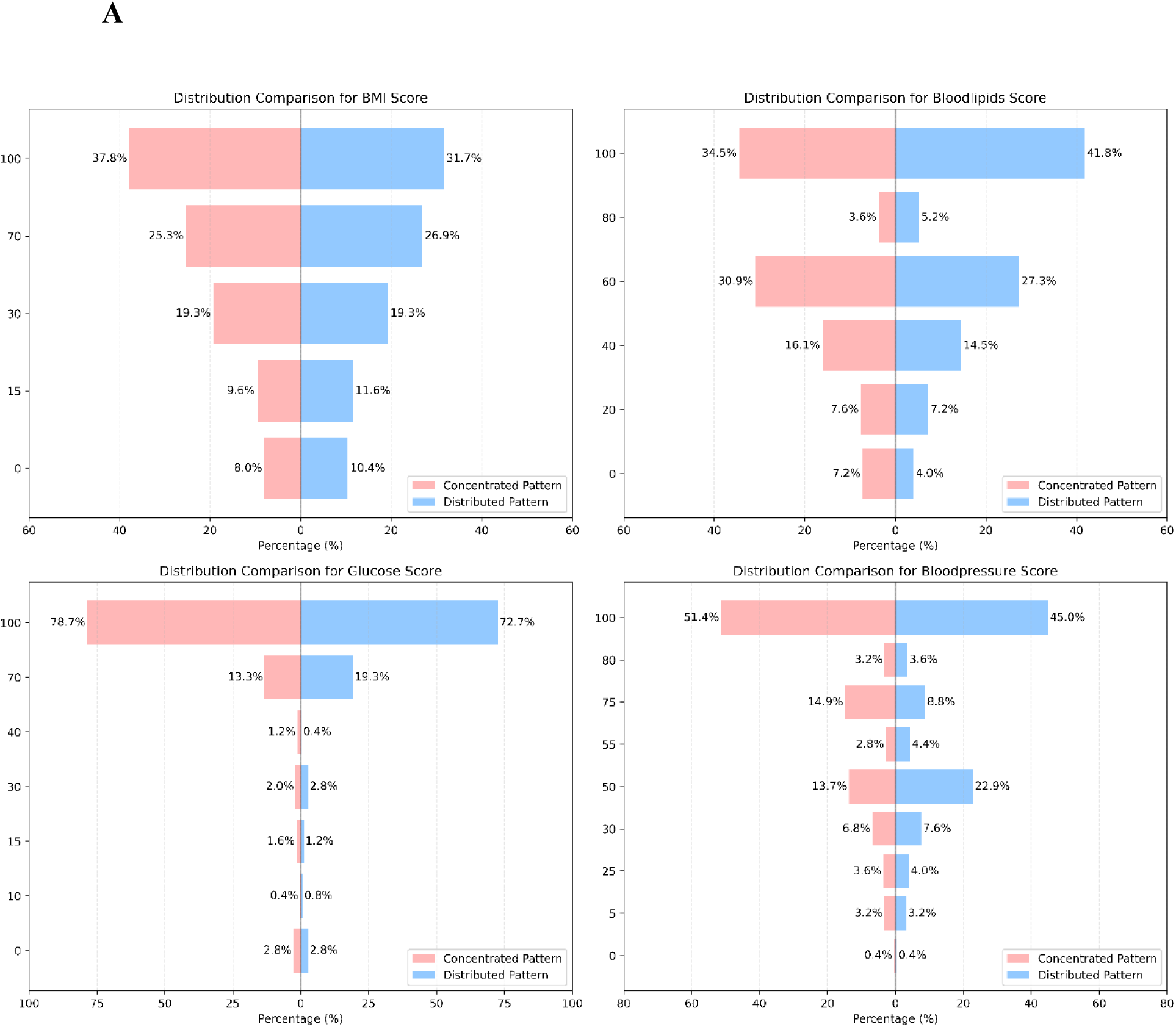
LE8 health factors by weekly accumulation pattern in low-volume participants. Tornado diagrams comparing BMI, blood lipids, glucose, and blood pressure scores between distributed and concentrated groups after matching. Concentrated patterns were associated with higher blood pressure scores.

**Table 3A.** Characteristics of low-volume participants before and after matching.

|  | All Low-Volume PA Participants |  |  | PSM Participants |  |  |
| --- | --- | --- | --- | --- | --- | --- |
|  | Unmatched<br>Concentrated (n =<br>471) | Unmatched<br>Distributed (n =<br>317) | Unmatched<br>\|SMD\| | Matched<br>Concentrated (n =<br>249) | Matched<br>Distributed (n =<br>249) | Matched<br>\|SMD\| |
| Female, n (%) | 232 (49.26%) | 203(64.04%) | 0.3466 | 155 (62.24%) | 153 (61.64%) | 0.0083 |
| Age, median (IQR) | 39 (29-50) | 45(35 - 55) | 0.4152 | 43(30-53) | 42(33-52) | 0.0354 |
| Race |  |  | 0.0567 |  |  | 0.0407 |
| Non-Hispanic Asian, n (%) | 55 (11.68%) | 42 (13.25%) |  | 29 (11.65%) | 31 (12.45%) |  |
| Non-Hispanic Black, n (%) | 84 (17.83%) | 62 (19.56%) |  | 45 (18.07%) | 49 (19.68%) |  |
| Non-Hispanic White, n (%) | 193 (40.98%) | 111 (35.02%) |  | 100 (40.16%) | 94 (37.75%) |  |
| Mexican American, n (%) | 63 (13.38%) | 51 (16.09%) |  | 37 (14.86%) | 43 (17.27%) |  |
| Other Hispanic, n (%) | 48 (10.19%) | 31 (9.78%) |  | 25 (10.04%) | 21 (8.43%) |  |
| Other races and ethnicity, n (%) | 28 (5.94%) | 20 (6.31%) |  | 13 (5.22%) | 11 (4.42%) |  |
| Education |  |  | 0.1351 |  |  | 0.0345 |
| High school or less | 162 (34.39%) | 115 (36.28%) |  | 91 (36.55%) | 87 (34.94%) |  |
| Some college | 141 (29.94%) | 114 (35.96%) |  | 89 (35.74%) | 87 (34.94%) |  |
| College Graduate | 168 (35.67%) | 87 (27.44%) |  | 69 (27.71%) | 75 (31.33%) |  |
| Family Income Ratio | 2.75(1.37-4.95) | 2.55(1.16-4.62) | 0.0942 | 2.56(1.20-4.59) | 2.68(1.18-4.61) | 0.0394 |
| Diet Score, median (IQR) | 50 (25 - 80) | 50 (25-50) | 0.1432 | 50 (25 - 80) | 25(25-50) | 0.0410 |
| Nicotine Score, median (IQR) | 100 (65-100) | 100 (60-100) | 0.0746 | 100 (50-100) | 100 (55-100) | 0.0071 |
| Sleep Score, median (IQR) | 100 (70 -100) | 100 (70-100) | 0.1266 | 100 (70-100) | 100 (70-100) | 0.0292 |

- Values are median (IQR) or n (%). Adequate covariate balance was defined as |SMD| <0.10.

**Table 4A.** Comparison of LE8 Health Factors Between Concentrated and Distributed Weekly Accumulation Patterns in the Low- Volume PA group After PSM.

|  | PS-all participants<br>(n = 498) | Concentrated Group<br>(n = 249) | Distributed Group<br>(n = 249) | P value |
| --- | --- | --- | --- | --- |
| BMI score, median (IQR) | 70 (30-100) | 70 (30-100) | 70 (30-100) | 0.142 |
| Blood lipids Score, median (IQR) | 60 (40-100) | 60(40-100) | 60(40-100) | 0.053 |
| Blood glucose Score, median (IQR) | 100 (100-100) | 100 (100-100) | 100 (60-100) | 0.325 |
| Blood pressure Score, median (IQR) | 80 (50-100) | 100 (50-100) | 75 (50-100) | 0.045 |

#### Intermediate-Volume Stratum

Among 994 intermediate-volume participants, 387 matched pairs were identified after PSM with excellent covariate balance (*Table 3B*). Concentrated patterns (≥45 minutes on active days) were associated with more favorable lipid (median 80 vs. 60; p=0.013) and glucose scores (p<0.001), with a modest improvement in BMI (p=0.044). Blood pressure did not differ between groups (*Table 4B, Figure 3B*).

**Figure 3B.**
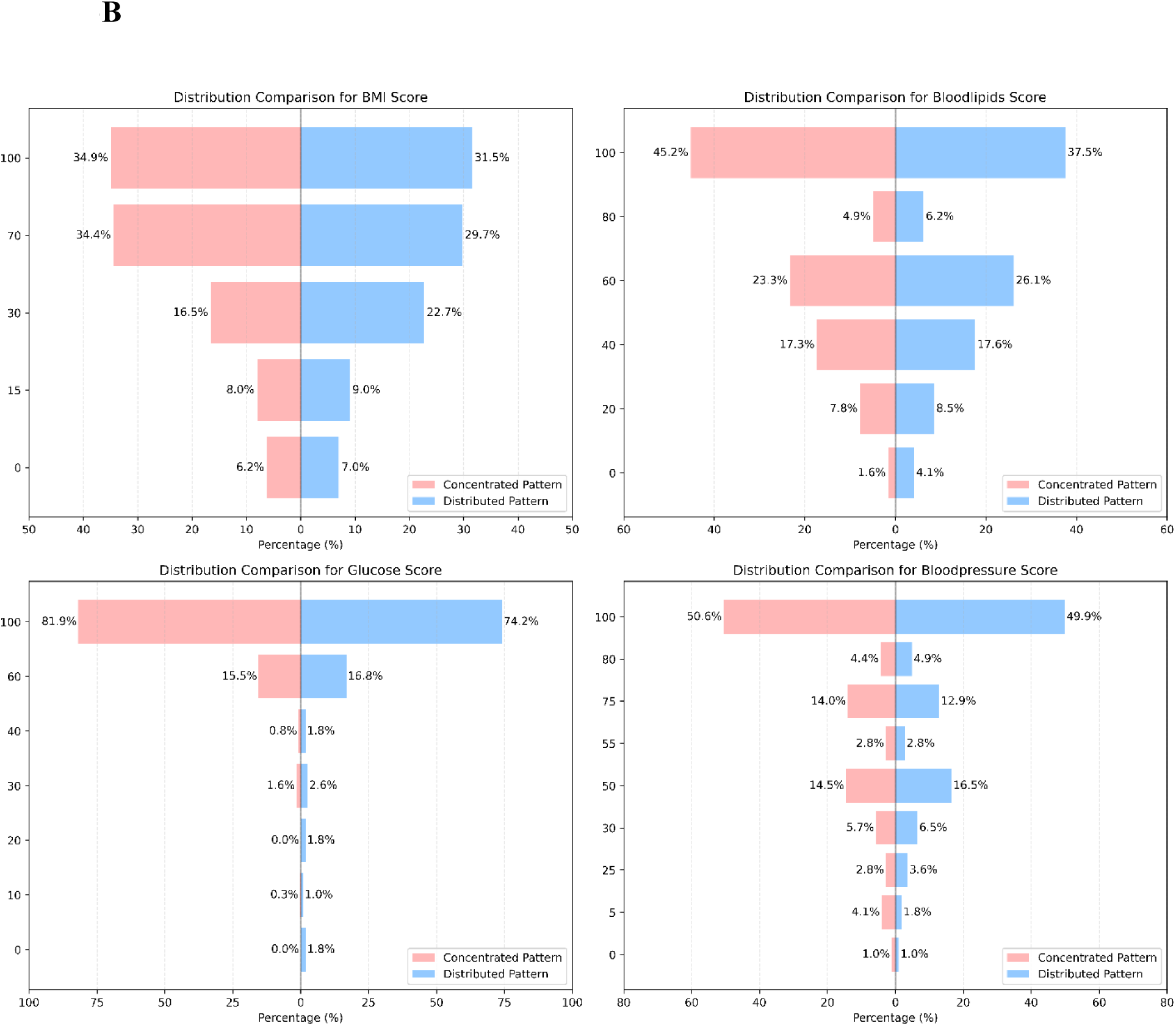
LE8 health factors by weekly accumulation pattern in intermediate-volume participants. Tornado diagrams comparing BMI, blood lipids, glucose, and blood pressure scores between distributed and concentrated groups after matching. Concentrated groups were associated with better blood lipid, and glucose scores, with no difference in blood pressure.

**Table 3B.**
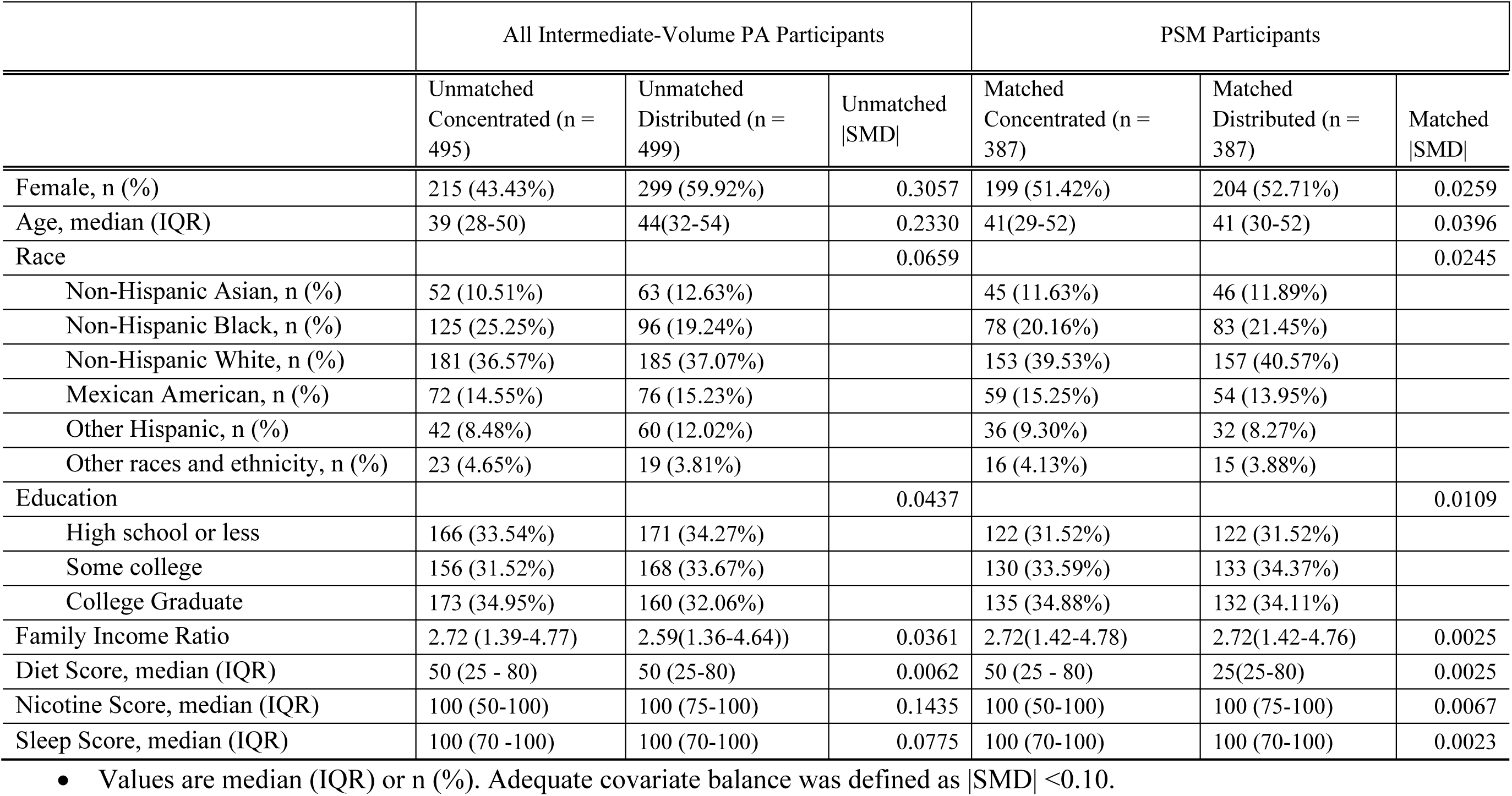
Characteristics of intermediate-volume participants before and after matching.

**Table 4B.** Comparison of LE8 Health Factors Between Concentrated and Distributed Weekly Accumulation Patterns in the Intermediate-Volume PA group After PSM.

|  | PS-all participants<br>(n = 774) | Concentrated Group<br>(n = 387) | Distributed Group<br>(n = 387) | P value |
| --- | --- | --- | --- | --- |
| BMI score, median (IQR) | 70 (30-100) | 70 (30-100) | 70 (30-100) | 0.044 |
| Blood lipids Score, median (IQR) | 60 (40-100) | 80 (40-100) | 60 (40-100) | 0.013 |
| Blood glucose Score, median (IQR) | 100 (100-100) | 100 (100-100) | 100 (60-100) | <0.001 |
| Blood pressure Score, median (IQR) | 100 (50-100) | 100 (50-100) | 80 (50-100) | 0.995 |

## DISCUSSION

In this analytic cohort of US adults without established CVD, both total weekly PA and the duration of activity on active days were associated with CVH as measured by the AHA LE8 framework. Consistent with prior work, higher weekly PA volume was linked to progressive improvements across multiple health domains. Beyond confirming the established dose-response relationship with weekly PA volume, our study provides new evidence that the structure of activity across the week is independently relevant. Among low-volume exercisers, concentrated patterns of ≥ 30 minutes on active days were associated with improved blood pressure, while in intermediate-volume exercisers, ≥ 45 minutes were linked with more favorable glucose and lipid regulation. These findings suggest that not only how much activity is performed, but also how it is distributed across the week, may influence cardiovascular prevention.

### Comparison with Prior Literature

Decades of epidemiologic research have demonstrated graded inverse associations between PA volume and cardiovascular morbidity and mortality.^31,32^ Reflecting this evidence, current recommendations emphasize weekly totals, with little attention to how activity is organized.

Studies that do address structure have largely focused on whether activity should be performed in long continuous sessions or divided into shorter bouts within a day. Trials in this area typically compared regimens such as three 10-minute bouts or two 15-minute bouts with a single 30-minute session, usually totaling 90-150 minutes per week.^33,34^ A meta-analysis of 19 randomized and quasi-experimental trials involving 1,080 adults concluded that continuous and accumulated bouts within a day produced broadly similar improvements in cardiorespiratory fitness, blood pressure, and most blood biomarkers, with some evidence that accumulated activity may be more favorable for weight and LDL cholesterol.^35^ However, these interventions were generally short-term (4-20 weeks), rarely tested sustained sessions beyond 40 minutes, and, most importantly, focused on within-day bout structure rather than the distribution of activity across the week.

Our study addresses this gap by evaluating weekly accumulation patterns—specifically, whether adults with suboptimal weekly totals concentrated activity into longer durations on fewer active days or distributed shorter durations across more days. This construct has not been incorporated into guidelines and remains underexplored in the literature. By leveraging nationally representative NHANES data, our analysis reflects habitual, long-term behaviors rather than prescribed interventions, and by applying the AHA LE8 framework, we captured both behavioral and biological dimensions of cardiovascular health. Within this framework, weekly accumulation patterns were independently associated with blood pressure, glucose, and lipid profiles, even among adults performing below guideline levels.

Mechanistic studies provide additional support. Sustained activity on active days has been shown to elicit threshold-dependent adaptations in vascular tone, glucose uptake, and mitochondrial function that fragmented activity may not fully trigger.^36,37^ Thus, these biological insights and our population-based findings suggest that while weekly structure may not substantially alter outcomes once guideline-level PA is achieved, accumulating ≥ 30-45 minutes on active days provides added benefits for adults with suboptimal weekly activity. Together, these findings extend prior work by moving beyond the question of "long versus short bouts in a single day" to demonstrate that the distribution of activity across the week is an independent determinant of cardiovascular health—a dimension not currently addressed in existing literature.

### Mechanistic Considerations

Several biological mechanisms may explain why active-day duration influences CVH. For blood pressure, aerobic activity performed for ≥30 minutes on exercise days produces post-exercise hypotension lasting up to 24 hours, mediated by reductions in sympathetic tone and enhanced nitric oxide-dependent vasodilation.^38,39^ This may underline the more substantial blood pressure benefits observed among low-volume participants exercising ≥30 minutes on active days.

For glucose regulation, continuous activity of at least 20-30 minutes enhances GLUT4 translocation, glycogen depletion, and mitochondrial biogenesis – processes that help explain why ≥45 minutes on active days were more strongly associated with glycemic improvements in intermediate-volume participants.^40,41^

For lipid metabolism, longer active-day durations may trigger threshold-dependent upregulation of lipoprotein lipase, enhancing triglyceride clearance and improving HDL function.^42^ Such effects are less likely to be elicited by fragmented activity performed in short durations.

Additional mechanisms—including improvements in vascular compliance, autonomic regulation^39^, and circadian influences on cardiometabolic responses^43^—further support the concept that the way activity is distributed across active days, independent of total weekly volume, can modify cardiovascular and metabolic health.

### Clinical and Public Health Implications

Our findings suggest that how adults structure their available activity time is an important determinant of overall CVH, as captured by the AHA LE8. For the large proportion of US adults who fall short of guidance-recommended PA volumes, accumulating at least 30-45 minutes on active days was associated with measurable benefits in blood pressure, glucose, and lipid metrics—core components of the LE8 framework. These results indicate that weekly accumulation patterns represent a practical and underrecognized level for optimizing CVH, even in individuals with limited weekly activity.

From a clinical standpoint, this shifts the conversation from "do more" toward "structure what you can do." People with limited time can still make meaningful gains by concentrating activity into long enough durations on active days, without necessarily increasing their total weekly minutes. This time-neutral approach aligns with precision-prevention efforts and provides clinicians with an actionable way to personalize exercise counseling within the broader context of CVH promotion.

At the public health level, our results also speak directly to the "weekend warrior" model, in which adults concentrate most of their activity into one or two days per week.^44^ While current guidelines encourage accumulating activity in any duration,^34^ our findings extend this framework by showing that when feasible, concentrated activity on active days delivers added benefit across multiple domains of CVH. This perspective broadens opportunities for equitable prevention, particularly for working adults and populations facing structural barriers to daily activity.

Finally, the identified active-day thresholds overlap with those needed to improve cardiorespiratory fitness (CRF), a powerful determinant of long-term cardiovascular outcomes.^45^ Thus, advising patients not only on how much to move but also on how to structure their activity across the week may enhance both short-term CVH metrics and long-term fitness trajectories, strengthening preventive strategies across the lifespan.

### Limitations and Future Directions

This study has several limitations. First, our analyses were restricted to an analytic cohort and did not incorporate survey weights; therefore, they should not be interpreted as nationally representative estimates. Second, the cross-sectional design precludes causal inferences; although PSM reduced measured confounding, residual bias from unmeasured factors remains possible.^22^ Third, PA was self-reported, introducing potential recall and reporting bias; objective assessment with accelerometry or wearable devices would strengthen future research. Fourth, outcomes were LE8 component scores rather than hard clinical endpoints, and subgroup analyses – particularly at the extremes of active-day duration – were limited by sample size. Finally, excluding the 2021-2023 cycle avoided pandemic-related confounding but may reduce the contemporary generalizability of the findings.

Future studies should prospectively evaluate active-day duration using objective monitoring and link weekly accumulation patterns to incident CVD and mortality. Randomized trials directly comparing concentrated versus distributed weekly accumulation at equivalent weekly volumes would provide mechanistic and clinical confirmation. Exploration of heterogeneity by age, sex, race/ethnicity, and baseline risk may help identify groups most likely to benefit from concentrated activity. In addition, integrating active-day duration targets into digital health platforms and community-based interventions could improve adoption, particularly among adults with limited time for exercise.

## CONCLUSION

In this analytic cohort of US adults without established CVD, accumulating ≥30-45 minutes on active days was associated with more favorable LE8 scores, even when total weekly activity remained below guideline targets. These findings highlight that how activity is distributed across the week—not only how much is accumulated—can meaningfully influence CVH. Recognizing active-day duration as a practical, time-efficient, and modifiable factor has important clinical and public health implications, particularly for adults with limited time who struggle to meet weekly activity recommendations.

## Data Availability

The data that support the findings of this study are publicly available from the National Health and Nutrition Examination Survey (NHANES) at the Centers for Disease Control and Prevention (CDC).

https://www.cdc.gov/nchs/nhanes/

## ABBREVIATIONS

AHA: American Heart Association
CVD: Cardiovascular Disease
CVH: Cardiovascular Health
IQR: Interquartile Range
LE8: Life’s Essential 8
LOESS: Locally Weighted Scatterplot Smoothing
NHANES: National Health and Nutrition Examination Survey
PA: Physical Activity
PSM: Propensity Score Matching

## ACKNOWLEDGMENTS

None.

## SOURCES OF FUNDING

None.

## DISCLOSURES

None.

## Notes

### Competing Interest Statement

The authors have declared no competing interest.

### Funding Statement

No external funding was received.

### Author Declarations

This study was determined exempt by the Institutional Review Board of Maanshan General Hospital of Ranger-Duree Healthcare, as it involves secondary analysis of publicly available de-identified data (NHANES).

## REFERENCE

1. Lindstrom M, DeCleene N, Dorsey H, et al. Global Burden of Cardiovascular Diseases and Risks Collaboration, 1990-2021. J Am Coll Cardiol. 2022;80(25):2372-2425. doi:10.1016/j.jacc.2022.11.001

2. Martin SS, Aday AW, Almarzooq ZI, et al. 2024 Heart Disease and Stroke Statistics: A Report of US and Global Data from the American Heart Association. Vol 149.; 2024. doi:10.1161/CIR.0000000000001209

3. Kazi DS, Elkind MSV, Deutsch A, et al. Forecasting the Economic Burden of Cardiovascular Disease and Stroke in the United States Through 2050: A Presidential Advisory From the American Heart Association. Circulation. 2024;150(4):e89–e101. doi:10.1161/CIR.0000000000001258

4. Slater K, Colyvas K, Taylor R, Collins CE, Hutchesson M. Primary and secondary cardiovascular disease prevention interventions targeting lifestyle risk factors in women: A systematic review and meta-analysis. Front Cardiovasc Med. 2022;9. doi:10.3389/fcvm.2022.1010528

5. Patnode CD, Redmond N, Iacocca MO, Henninger M. Behavioral Counseling Interventions to Promote a Healthy Diet and Physical Activity for Cardiovascular Disease Prevention in Adults Without Known Cardiovascular Disease Risk Factors: Updated Evidence Report and Systematic Review for the US Preventive Serv. Jama. 2022;328(4):375–388. doi:10.1001/jama.2022.7408

6. Tian D, Meng J. Exercise for prevention and relief of cardiovascular disease: Prognoses, mechanisms, and approaches. Oxid Med Cell Longev. 2019;2019(Mi). doi:10.1155/2019/3756750

7. Lloyd-Jones DM, Hong Y, Labarthe D, et al. Defining and setting national goals for cardiovascular health promotion and disease reduction: The american heart association’s strategic impact goal through 2020 and beyond. Circulation. 2010;121(4):586–613. doi:10.1161/CIRCULATIONAHA.109.192703

8. Lloyd-Jones DM, Allen NB, Anderson CAM, et al. Life’s Essential 8: Updating and Enhancing the American Heart Association’s Construct of Cardiovascular Health: A Presidential Advisory from the American Heart Association. Circulation. 2022;146(5):E18–E43. doi:10.1161/CIR.0000000000001078

9. Arnett DK, Blumenthal RS, Albert MA, et al. 2019 ACC/AHA Guideline on the Primary Prevention of Cardiovascular Disease: A Report of the American College of Cardiology/American Heart Association Task Force on Clinical Practice Guidelines. Vol 140.; 2019. doi:10.1161/CIR.0000000000000678

10. Bull FC, Al-Ansari SS, Biddle S, et al. World Health Organization 2020 guidelines on physical activity and sedentary behaviour. Br J Sports Med. 2020;54(24):1451-1462. doi:10.1136/bjsports-2020-102955

11. Cheng W, Zhang Z, Cheng W, Yang C, Diao L, Liu W. Associations of leisure-time physical activity with cardiovascular mortality: A systematic review and meta-analysis of 44 prospective cohort studies. Eur J Prev Cardiol. 2018;25(17):1864–1872. doi:10.1177/2047487318795194

12. Lear SA, Hu W, Rangarajan S, et al. The effect of physical activity on mortality and cardiovascular disease in 130 000 people from 17 high-income, middle-income, and low-income countries: the PURE study. Lancet. 2017;390(10113):2643–2654. doi:10.1016/S0140-6736(17)31634-3

13. MacDonald JR, MacDougall JD, Hogben CD. The effects of exercise duration on post-exercise hypotension. J Hum Hypertens. 2000;14(2):125–129. doi:10.1038/sj.jhh.1000953

14. Halliwill JR, Buck TM, Lacewell AN, Romero SA. Postexercise hypotension and sustained postexercise vasodilatation: What happens after we exercise? Exp Physiol. 2013;98(1):7–18. doi:10.1113/expphysiol.2011.058065

15. Sylow L, Kleinert M, Richter EA, Jensen TE. Exercise-stimulated glucose uptake-regulation and implications for glycaemic control. Nat Rev Endocrinol. 2017;13(3):133–148. doi:10.1038/nrendo.2016.162

16. Way KL, Hackett DA, Baker MK, Johnson NA. The effect of regular exercise on insulin sensitivity in type 2 diabetes mellitus: A systematic review and meta-analysis. Diabetes Metab J. 2016;40(4):253–271. doi:10.4093/dmj.2016.40.4.253

17. Stockwell S, Trott M, Tully M, et al. Changes in physical activity and sedentary behaviours from before to during the COVID-19 pandemic lockdown: A systematic review. BMJ Open Sport Exerc Med. 2021;7(1):1–8. doi:10.1136/bmjsem-2020-000960

18. Lloyd-Jones DM, Ning H, Labarthe D, et al. Status of Cardiovascular Health in US Adults and Children Using the American Heart Association’s New "Life’s Essential 8" Metrics: Prevalence Estimates From the National Health and Nutrition Examination Survey (NHANES), 2013 Through 2018. Circulation. 2022;146(11):822-835. doi:10.1161/CIRCULATIONAHA.122.060911

19. Mellen PB, Gao SK, Vitolins MZ, Goff DC. Deteriorating Dietary Habits Among Adults With Hypertension. Arch Intern Med. 2008;168(3):308. doi:10.1001/archinternmed.2007.119

20. Jones AE. Lactate Clearance vs Central Venous Oxygen Saturation as Goals of Early Sepsis Therapy <subtitle>A Randomized Clinical Trial</subtitle>. JAMA. 2010;303(8):739. doi:10.1001/jama.2010.158

21. Gijbels I, Prosdocimi I. Loess. Wiley Interdiscip Rev Comput Stat. 2010;2(5):590–599. doi:10.1002/wics.104

22. Austin PC. An introduction to propensity score methods for reducing the effects of confounding in observational studies. Multivariate Behav Res. 2011;46(3):399–424. doi:10.1080/00273171.2011.568786

23. Rosenbaum PR, Rubin DB. The central role of the propensity score in observational studies for causal effects. Matched Sampl Causal Eff. 2006;(1083):170–184. doi:10.1017/CBO9780511810725.016

24. Rosenbaum PR, Rubin DB. Reducing Bias in Observational Studies Using Subclassification on the Propensity Score. J Am Stat Assoc. 1984;79(387):516–524. doi:10.1080/01621459.1984.10478078

25. Austin PC. Balance diagnostics for comparing the distribution of baseline covariates between treatment groups in propensity-score matched samples. Stat Med. 2009;28(25):3083–3107. doi:10.1002/sim.3697

26. Stuart A. A TEST FOR HOMOGENEITY OF THE MARGINAL DISTRIBUTIONS IN A TWO-WAY CLASSIFICATION. Biometrika. 1955;42(3-4):412–416. doi:10.1093/biomet/42.3-4.412

27. Maxwell AE. Comparing the Classification of Subjects by Two Independent Judges. Br J Psychiatry. 1970;116(535):651–655. doi:10.1192/bjp.116.535.651

28. Chen TC, Parker JD, Clark J, Shin HC, Rammon JR, Burt VL. National Health and Nutrition Examination Survey: Estimation Procedures, 2011-2014. Vital Health Stat 2. 2018;(177):1-26.

29. Health N, Survey E. Vital and Health Statistics National Health and Nutrition Examination Survey, 2017 – March 2020 Prepandemic File: Sample Design, Estimation, and Analytic Guidelines Data Evaluation and Methods Research. 2022;(190).

30. Chen TC, Clark J, Riddles MK, Mohadjer LK, Fakhouri THI. National Health and Nutrition Examination Survey, 2015-2018: Sample Design and Estimation Procedures. Vital Health Stat 2. 2020;(184):1-35.

31. Kazemi A, Soltani S, Aune D, et al. Leisure-time and occupational physical activity and risk of cardiovascular disease incidence: a systematic-review and dose-response meta-analysis of prospective cohort studies. Int J Behav Nutr Phys Act. 2024;21(1):1–15. doi:10.1186/s12966-024-01593-8

32. Lear SA, Hu W, Rangarajan S, et al. The effect of physical activity on mortality and cardiovascular disease in 130 000 people from 17 high-income, middle-income, and low-income countries: the PURE study. Lancet. 2017;390(10113):2643–2654. doi:10.1016/S0140-6736(17)31634-3

33. Saint-Maurice PF, Troiano RP, Bassett DR, et al. Association of Daily Step Count and Step Intensity With Mortality Among US Adults. JAMA. 2020;323(12):1151. doi:10.1001/jama.2020.1382

34. Piercy KL, Troiano RP, Ballard RM, et al. The Physical Activity Guidelines for Americans. JAMA. 2018;320(19):2020. doi:10.1001/jama.2018.14854

35. Murphy MH, Lahart I, Carlin A, Murtagh E. The Effects of Continuous Compared to Accumulated Exercise on Health: A Meta-Analytic Review. Sport Med. 2019;49(10):1585–1607. doi:10.1007/s40279-019-01145-2

36. Kirwan JP, Sacks J, Nieuwoudt S. The essential role of exercise in the management of type 2 diabetes. Cleve Clin J Med. 2017;84(7):S15–S21. doi:10.3949/ccjm.84.s1.03

37. Karstoft K, Winding K, Knudsen SH, et al. The Effects of Free-Living Interval-Walking Training on Glycemic Control, Body Composition, and Physical Fitness in Type 2 Diabetic Patients. Diabetes Care. 2013;36(2):228-236. doi:10.2337/dc12-0658

38. Halliwill JR. Mechanisms and Clinical Implications of Post-exercise Hypotension in Humans. Exerc Sport Sci Rev. 2001;29(2):65–70. doi:10.1249/00003677-200104000-00005

39. Fisher JP, Young CN, Fadel PJ. Central sympathetic overactivity: Maladies and mechanisms. Auton Neurosci. 2009;148(1-2):5–15. doi:10.1016/j.autneu.2009.02.003

40. Richter EA, Hargreaves M. Exercise, GLUT4, and skeletal muscle glucose uptake. Physiol Rev. 2013;93(3):993–1017. doi:10.1152/physrev.00038.2012

41. Holloszy JO. Exercise-induced increase in muscle insulin sensitivity. J Appl Physiol. 2005;99(1):338–343. doi:10.1152/japplphysiol.00123.2005

42. Kiens B. Skeletal Muscle Lipid Metabolism in Exercise and Insulin Resistance. Physiol Rev. 2006;86(1):205–243. doi:10.1152/physrev.00023.2004

43. Sato S, Basse AL, Schönke M, et al. Time of Exercise Specifies the Impact on Muscle Metabolic Pathways and Systemic Energy Homeostasis. Cell Metab. 2019;30(1):92–110.e4. doi:10.1016/j.cmet.2019.03.013

44. O’Donovan G, Lee IM, Hamer M, Stamatakis E. Association of "Weekend Warrior" and Other Leisure Time Physical Activity Patterns With Risks for All-Cause, Cardiovascular Disease, and Cancer Mortality. JAMA Intern Med. 2017;177(3):335. doi:10.1001/jamainternmed.2016.8014

45. Ross R, Blair SN, Arena R, et al. Importance of Assessing Cardiorespiratory Fitness in Clinical Practice: A Case for Fitness as a Clinical Vital Sign: A Scientific Statement From the American Heart Association. Circulation. 2016;134(24). doi:10.1161/CIR.0000000000000461

